# Behavioral and Functional Neuroimaging Effects of Delivering a Course of Repetitive Transcranial Magnetic Stimulation to Personalized Targets Within the Ventrolateral Or Dorsolateral Prefrontal Cortex in Treatment-Seeking Participants with Cannabis Use Disorder

**DOI:** 10.64898/2026.06.08.26355193

**Authors:** Daniel McCalley, Brendan Wong, Andrew Geoly, Wiebke Struckman, Azeezat Azeez, Irakli Kaloiani, Bohye Kim, Seigo Ninomiya, Jarrod Ehrie, Christopher W. Austelle, Cameron E. Rolle, Jane P. Kim, Brett Froeliger, Aimee L. McRae-Clark, Gregory L. Sahlem

**Author notes:** **Corresponding author:** Gregory L. Sahlem, M.D. Associate Professor, Duke University Department of Psychiatry and Behavioral Sciences 2400 Pratt St, Durham, NC, 27514. **Author Contributions:** BW, AG, WS, AA, BF, AM, and GS designed the experiment. BW, AG, WS, AA, IK, SN, JE, CA, CR, and GS conducted the experiment. DM, BW, BK, JK, and GS performed the analysis. DM, AG, BK, WS, and GS wrote the first draft of the manuscript. All authors critically reviewed and edited the manuscript. **Clinicaltrials.gov identifiers:** NCT05720312.

## Abstract

**Background:** Repetitive Transcranial Magnetic Stimulation (rTMS) is a promising treatment across addictive disorders including Cannabis Use Disorder (CUD). Stimulation of two rTMS-targets, the ventromedial prefrontal cortex (vmPFC) and the left dorsolateral prefrontal cortex (LDLPFC), limbic and executive control network hubs respectively, may yield differential effects. In this pilot trial, we explored the differential effects of 36-sessions of rTMS applied to either the vmPFC or LDLPFC.

**Methods:** Treatment-seeking participants with moderate or severe CUD (n=20, 10F, age=33.3±9.8SD) were randomized to 36-sessions of open-label rTMS (two sessions-per-visit, two or three visits-per-week) to either the LDLPFC (3000-pulses; 10Hz) or vmPFC (900-pulses; 1Hz) using personalized functional Magnetic Resonance Imaging (fMRI) targets along with three-sessions of Motivational Enhancement Therapy. At baseline and following rTMS, the Time-Line Follow-Back was used to measure Days-per-week of cannabis use and the fMRI Regulation of Craving (ROC) task was used to measure network activation to cues associated with long-term negative (‘Later’) and short-term positive (‘Now’) consequences of cannabis use.

**Results:** Eighty percent of participants completed study-rTMS. There was a significant decrease in days-per-week of cannabis use in both groups (vmPFC: d=7.9; DLPFC, d=3.1) between the four-weeks of baseline and seven-weeks of follow-up. LDPFC-rTMS reduced fMRI BOLD signal magnitude and increased LDLPFC functional connectivity in response to cues, while vmPFC-TMS reduced functional connectivity.

**Conclusions:** Treatment-seeking participants with CUD reduced the number of days-per-week they used cannabis when receiving rTMS applied to either the LDPFC or vmPFC, while fMRI effects differed by treatment target. Future larger sham-controlled trials are needed for efficacy and biomarker determination.

## Introduction

Cannabis Use Disorder (CUD) is a highly prevalent addictive disorder with demonstrated morbidity^1–4^. Behavioral treatment paradigms have consistently yielded moderate treatment effects, and several medications have shown promise in the treatment of CUD; however, no medication has emerged as clearly effective for CUD and long-term treatment responses overall remain low^5–8^. As such, there is a need for novel treatment development for CUD. Our group recently completed a Phase 2 trial that demonstrated the promise of high-frequency repetitive Transcranial Magnetic Stimulation (rTMS) applied to the Left Dorsolateral Prefrontal Cortex (LDLPFC)^9^ in the treatment of CUD. Though we found moderate treatment effects in a sham-controlled trial, we made several methodological compromises in this initial trial to improve retention, given a low retention rate in an earlier pilot trial^10^. The main methodologic compromises included a relatively low number of delivered sessions of rTMS (twenty total) and scalp-based(using Beam-F3^11^). We reasoned that this lower participant burden paradigm maximized feasibility while delivering enough rTMS to produce a therapeutic effect. While our treatment paradigm was feasible (70% retention rate), it is possible that refining these initial compromises would improve efficacy while maintaining feasibility.

In addition to these experimental refinements, another open-ended question in the field which may improve treatment efficacy is: *which cortical region should be targeted with rTMS?* There are two predominant approaches for rTMS in the addiction literature – targeting the DLPFC as a strategy to enhance executive control over use and targeting the vmPFC as a strategy to reduce limbic drive toward use^12^. Bolstering this conceptual framework, a single pulse of TMS applied to the DLPFC and vmPFC travel to different downstream targets in the dorsal and ventral striatum, respectively, indicating these cortical regions are hubs of discrete neural circuits^12,13^. Until now, both of these strategies have been evaluated separately in the literature, but very few studies have evaluated the relative influence of both strategies on reducing substance use. Thus, there is a gap in our knowledge regarding whether these treatment targets will yield the same or different effects on the brain and behaviors related to cannabis use.

In the present pilot trial, we preliminarily evaluated the influence of LDLPFC and vmPFC rTMS on changes in clinical and neuroimaging outcomes leveraging the following innovations to our experimental design: 1) <u>*36 sessions of rTMS*</u> and 2) <u>*individualized, fMRI-guided targeting*</u>. Both of these strategies have shown enhanced clinical benefit in improving rTMS response and remission rates among patients with depression and addiction ^13,14–16,17^. To advance the field’s understanding of the influence of these two treatment strategies on brain circuitry, we developed an fMRI based rTMS targeting paradigm to engage the LDPFC and vmPFC and their downstream afferent targets^18–20^. Specifically, we adapted the Regulation of Craving (ROC) fMRI task^18^ to generate rTMS treatment targets and to measure changes in cannabis cue reactivity^20^. This task is designed to present cues to individuals under two different contexts: considering the immediate, rewarding aspects of use (‘Now’ trials) to engage the vmPFC and incentive-salience circuitry and considering long-term, negative consequences (‘Later’ trials) of use to engage the LDLPFC and executive function circuitry.

We hypothesized that delivering our enhanced treatment paradigm would be feasible (pre-specifying a feasibility cut-off of 60% retention), and that both strategies would increase weeks of abstinence and reduce cannabis use days. We further hypothesized that participants would have improved quality of life at the end of a 6-week follow-up period relative to their baseline quality of life. Finally, we explored the differential effects of stimulating each rTMS-target on behavioral constructs and fMRI-based outcomes. Notably, to our knowledge, the vmPFC has never been selected as an rTMS target within a cannabis use disordered population. We specifically hypothesized that LDLPFC applied rTMS would affect LDLPFC activation in the LATER runs of the ROC-task, and that vmPFC-applied rTMS would affect vmPFC activation in the NOW runs of the ROC task.

## Methods

### Overview, Recruitment, Screening, Treatment, and Assessment

In this pre-registered clinical trial (NCT05720312), we recruited treatment-seeking participants with moderate or severe CUD through clinical referrals from the Stanford University Addiction Medicine clinic and from the area around Palo Alto, California using media advertisements. The trial was approved by the Stanford University Institutional Review Board, was conducted in accordance with the principles outlined in the Declaration of Helsinki, and all participants signed informed consent prior to beginning study procedures. The trial was split into three time periods, which included: a) a screening/baseline MRI period including assessment of cannabis use patterns for the 4-weeks prior to screening; b) an acute treatment period which included 18 rTMS visits over a period of six to nine weeks (two to three visits-per-week based on participant preference, 2 rTMS sessions/visit, three-sessions of Motivational Enhancement Therapy), and; c) a post treatment MRI and follow-up period which included a post-treatment MRI occurring up to a week after the last rTMS-session and brief two, four, and six week follow-up visits where cannabis use was assessed (Table 1).

**Table 1:** Experimental Design. All participants provided baseline measures of previous 28-day cannabis use (Time-Line Follow-Back, TLFB), Cannabis craving (three item Craving Scale, CS) and delay discounting. Baseline fMRI measured brain response to cannabis cues using the Regulation of Craving (ROC) task. Participants were randomized to receive 4-6 sessions of repetitive Transcranial Magnetic Stimulation (rTMS) per week for 6-9 weeks during the rTMS period [Note: above figure represents 3X rTMS visits/week, e.g. a total of 6 weeks]. Follow up fMRI was completed within 1 week of rTMS completion. Cannabis use, craving, and delay discounting were measured at 1, 2, 4 and 6-week follow-up visits.

|  | Screen | Baseline | TMS Period |  |  |  |  |  | Follow Up |  |  |  |
| --- | --- | --- | --- | --- | --- | --- | --- | --- | --- | --- | --- | --- |
|  |  |  | 1 | 2 | 3 | 4 | 5 | 6 | 1 | 2 | 4 | 6 |
| **Consent** | **X** |  |  |  |  |  |  |  |  |  |  |  |
| **rTMS** |  |  | **XXX** | **XXX** | **XXX** | **XXX** | **XXX** | **XXX** |  |  |  |  |
| **fMRI** |  | **X** |  |  |  |  |  |  | **X** |  |  |  |
| **Cannabis Use (TLFB)** | **X** | **X** |  |  |  |  |  |  | **X** | **X** | **X** | **X** |
| **Craving** |  | **X** |  |  |  |  |  |  | **X** | **X** | **X** | **X** |
| **Delay Discounting** |  | **X** |  |  |  |  |  |  | **X** | **X** | **X** | **X** |

### Screening and pre-rTMS period

Participants underwent a structured screening and enrollment visit which included a structured clinical history and exam and assessment using the Quick Structured Clinical Interview for the 5^th^ edition of the Diagnostic and Statistical Manual of Mental Disorders^21^. Participants further were evaluated using the Time-Line Follow-Back (TLFB)^22^ with a 28-day time-frame, the Marijuana Problem Scale (MPS)^23^, the Short form of the Quality of Life and Enjoyment Scale (Q-LES-Q-SF)^24^, the three item Craving Scale (CS)^25^, and the Cannabis Withdrawal Scale (CWS)^26^. Participants were enrolled in the study if they were older than 18 years old; they met criteria for moderate or severe cannabis use disorder; expressed interest in cannabis cessation or reduction, and; had sufficient intelligence and command of the English language to complete informed consent procedures and study related assessments. Participants were excluded if they were pregnant or breastfeeding; met criteria for any other moderate or severe substance use disorder (other than tobacco use disorder); had changes in psychoactive medications in the 4-weeks preceding enrollment; had a history of bipolar affective disorder, schizophrenia, dementia, active suicidal ideation, or a suicide attempt within the past 6-months; had any contraindications to rTMS^27^ or MRI^28^, or; had other unstable medical, neurologic, or psychiatric disorders.

Participants who enrolled in the trial underwent a baseline MRI visit following 24-hours of abstinence from cannabis and other substances verified by both self-report and saliva drug testing (12-panel now, Boynton Beach, Fl). Prior to the MRI visit they underwent standard assessments including cannabis withdrawal^26^, craving^25^, and delayed discounting^29^. Following training, participants then underwent scanning as outlined below.

### rTMS treatment period

After completing the pre-treatment fMRI, functional targets were generated as described in^20^, and participants were randomized to receive rTMS to either the LDPFC or vmPFC in an open-label fashion. Each participant underwent a total of 36-sessions of rTMS delivered as two-sessions per-visit (with approximately 45-minutes between sessions), that either occurred two or three times-per-week based on participant preference. Participants also received three sessions of Motivational Enhancement Therapy^23^ on approximately the second, fourth, and sixth visits for participants undergoing twice-weekly visits, and the second, fifth, and ninth visits for participants attending three-weekly visits. All counseling sessions occurred in the inter-rTMS-session interval between the first and second rTMS session.

We used a MagVenture MagPro X100 device and a cool-B65 A/P coil (MagVenture, Denmark) in active mode to deliver a total of 36-sessions of rTMS using neuronavigation (Localite, Sankt Augustin, Germany). We performed a resting motor threshold (rMT) determination at baseline and delivered stimulation at a goal of 120% rMT. The rTMS paradigms differed by treatment target. A high-frequency paradigm was used for the DLPFC target (10Hz, 5-seconds on, 10-seconds off, 3000-pulses), while a low-frequency paradigm was used for the vmPFC target (1Hz, continuous, 900-pulses). Both paradigms were matched on time in the chair (15-minutes). A standardized cannabis cue-paradigm was presented prior to the delivery of each rTMS-session as described in our previous trial^9^, and participants were instructed to continuously think about a pleasurable cannabis use time (to induce cued-craving to directly suppress via vmPFC-rTMS or indirectly suppress via DLPFC-rTMS).

### Post-rTMS and Follow-up period

Following the final rTMS visit participants were scheduled for a post-rTMS MRI visit which occurred as soon as the following day and as long as 7-days later. Participants were again asked to refrain from using alcohol, cannabis or other substances (verified using self-report and saliva drug testing). Craving, withdrawal, and delayed discounting were assessed, again, as was urine drug testing, and the TLFB for the week preceding the MRI visit. Brief, in-person, follow-up visits were then scheduled two, four, and six weeks later, and occurred with ad libitum cannabis use for participants who still used cannabis. Craving, withdrawal, delayed-discounting, objective cannabis use (urine drug-testing), and subjective cannabis use (TLFB) were assessed at each of these visits.

### Behavioral data analysis

This exploratory trial had three main overarching goals, which included a) determining whether the enhanced treatment paradigm was feasible, b) determining preliminary treatment efficacy for either or both of the treatment targets, and c) determining whether there were suggestions of distinct treatment or imaging effects based on treatment target (LDLPFC and vmPFC). Given this was a proof-of-concept trial, we pre-specified that we would focus on effect sizes, and report *p*-values as pertinent only.

For feasibility, we pre-specified retention as the variable of interest and set a cut-off of 60%. We further measured the Perceived Research Burden Assessment (PeRBA), which has a score range from 22-110 with lower scores suggesting lower burden^30^, following the administration of 16-sessions of rTMS and 36-sessions of rTMS, to determine if there was a change in perceived burden. We used a within-subject t-test to test whether perceived research burden increased between lower and higher rTMS doses.

For treatment efficacy, we pre-specified self-reported weeks of abstinence following the administration of 16-sessions of rTMS (primary), days-per-week and sessions-per-week of cannabis use in the follow-up period, and the four-week continuous quit rate (any point in the study) as markers of efficacy. We imputed missing data as non-abstinent but left it missing for continuous measures (e.g. days-per-week). Given the small sample-size we elected to simply report the percent of weeks of abstinence and the percent of participants achieving four consecutive weeks of abstinence. To examine whether days-per-week of cannabis use and cannabis use-sessions-per-week changed from pre-treatment to follow-up in each treatment type, we recorded 4 weeks of pre-treatment timeframe as “pre-treatment” (i.e., 0) and 13 weeks of follow-up timeframe (last 6 weeks of rTMS and last 7 weeks of follow-up) as “follow-up” (i.e., 1) and fit Poisson mixed effects models or negative binomial mixed effects models as appropriate (i.e., depending on whether overdispersion was present or not) using a log link function for each treatment type. We generated effect sizes (i.e., Cohen’s D) based on the model estimates. We additionally pre-specified improvement of quality of life between the baseline and the 6-week follow-up as a secondary measure of treatment efficacy. To test the pre-to-post treatment effects in quality of life, we performed a paired t-test between the screening visit and the 6-week follow-up.

Finally, in an exploratory fashion we examined whether there were differential effects of vmPFC relative to DLPFC rTMS (i.e., group effects) on the three-item craving scale and the $1000 delayed discounting k-variable. We fit linear mixed effects models with time point, treatment type, and their interaction and evaluated significance of the interaction term.

### Neuroimaging data analysis

#### MR scanning parameters

MRI scans were completed on a GE Discovery MR750 scanner (GE Medical systems, Chicago, Illinois, USA) with a Nova 32-channel head coil. Functional images were acquired with a T2*-weighted EPI BOLD sequence with a TR of 1,600ms, TE of 30ms, flip angle of 60°, 116 x 116 in-plane matrix, field of view of 23.2cm, and 72 2mm-slices in an interleaved order. Anatomical images were acquired with a T1-weighted sequence with a TR of 6.388ms, TE of 2.624ms, flip angle of 12°, 256 x 256 in-plane matrix, field of view of 23.0cm, and 0.9mm-slices covering the entire brain of the participant. Phase Reverse scans were acquired for distortion correction.

#### fMRI Regulation of Craving (ROC) task

The MRI paradigm including our pre-processing and first level processing procedures, is described in detail in our previous targeting manuscript^20^. All MRI visits occurred following 24-hours of cannabis abstinence. Participants underwent a structured training session for the ROC-task prior to scanning.

#### fMRI data preprocessing

All functional data were preprocessed using standard methods in fMRIprep version 25.0.0^31,32^. Briefly, anatomical preprocessing included bias field correction, skull stripping, tissue segmentation, surface reconstruction, and nonlinear normalization to MNI standard space. Functional preprocessing involved motion correction, susceptibility distortion correction, co-registration to anatomical images, and spatial smoothing (6mm FWHM). Noise reduction was performed using ICA-AROMA for motion artefacts and CompCor for physiological noise, with confound regressors including framewise displacement (>0.5mm), DVRAS, and global signals. Preprocessed BOLD data were resampled into standard space using Lanczos interpolation.

#### First-level generalized linear modeling; BOLD signal magnitude and functional connectivity

General linear models (GLM) were constructed using the Statistical Parametric Mapping (SPM12, BOLD signal magnitude analyses) and the Connectivity toolboxes (Conn, connectivity analyses) implemented in MATLAB (R2018a). In both toolboxes, preprocessed, native-space, de-skulled, and grey-matter masked BOLD EPI data were input into each model. The model was fit with 11 task regressors (‘Now’, 3 regressors: prompt, image cue, and relax; ‘Later’, 3 regressors: prompt, image cue, and relax; Hand-pad ratings, 3 regressors: following now cue, following later cue, and following neutral cues; Fixation crosses, 2 regressors), 6 motion regressors (X, Y, Z, pitch, roll and yaw), a constant regressor, and a high pass filter cutoff of 100 seconds. Motion-adjusted contrast maps were created for each task condition. Primary contrasts of interest for between group analyses were ‘Now’ and ‘Later’ image cue responses.

#### Region-of-interest approach

Given the relatively small sample size in each group, we chose to evaluate change in cannabis cue-reactivity within specific regions of interest (ROI). Consistent with prior work, a priori network of regions were selected for analysis ^33,34^, which include: the stimulation targets, vmPFC and DLPFC, the bilateral anterior cingulate cortex, the dorsal striatum, the ventral striatum, and the left and right insula. Average BOLD signal during ‘Now’ and ‘Later’ cues was extracted for each ROI to measure relative BOLD signal magnitude (expressed as parameter estimate *β*). ROI-to-ROI weighted connectivity values were calculated from the stimulation target and the same set of brain regions (Fischer’s transformed correlation coefficient, z scores). The seed ROI for connectivity analyses were matched to the stimulation type received (e.g. LDPFC seed for LDLPFC stimulation, vmPFC seed for vmPFC stimulation).

#### Statistical Analysis of ROI data

To assess the change in BOLD signal magnitude and functional connectivity following each rTMS approach, univariate general linear models were constructed using IBM SPSS Statistics (version 26; IBM Corp.). Fixed factors included treatment type (LDLPFC or vmPFC rTMS), time (pre-TMS, post-TMS), and ROI. Hedge’s g was calculated to express the degree of change in BOLD signal magnitude and functional connectivity in the same units and to facilitate future clinical trial design. Last, in an exploratory analysis, we also evaluated whole-brain, voxel-wise change in BOLD signal magnitude in brain response to Now and Later cues (Figure-4).

## Results

### Participant Characteristics and feasibility (Table 2, Supplemental Figure 1)

We assessed a total of 30 potential participants for eligibility, of which 22 were eligible for enrollment. Twenty participants attended an initial rTMS visit and were randomized to DLPFC or vmPFC rTMS. Eight participants in each arm completed study-rTMS and post-rTMS MRI visits (for a pre-specified retention rate of 80%). Two participants were lost to follow-up in the DLPFC arm following the post-rTMS-MRI, and subsequently full data was available for 60% of the DLPFC group and 80% of the vmPFC group. PeRBA scores remained stable and low between assessments with a score of 32.8±8.3 following the administration of 16 sessions of rTMS and 33.0±8.0 following the administration of 36 sessions of rTMS. The study population consisted of 50% female participants, had a mean age of 33.3±9.8, and all met criteria for severe CUD. The sample was diverse with no racial/ethnic category exceeding 30% of the total. Given the small sample, we did not test for statistical significance between groups, but noted numerical differences suggesting the DLPFC group had more severe illness, including a higher proportion using concentrated cannabis, more use-sessions-per-day, and a generally lower cannabis reduction goal (6/10 participants in the vmPFC group stated a goal of abstinence, while only 20% of the DLPFC group had the same goal).

**Table 2:**
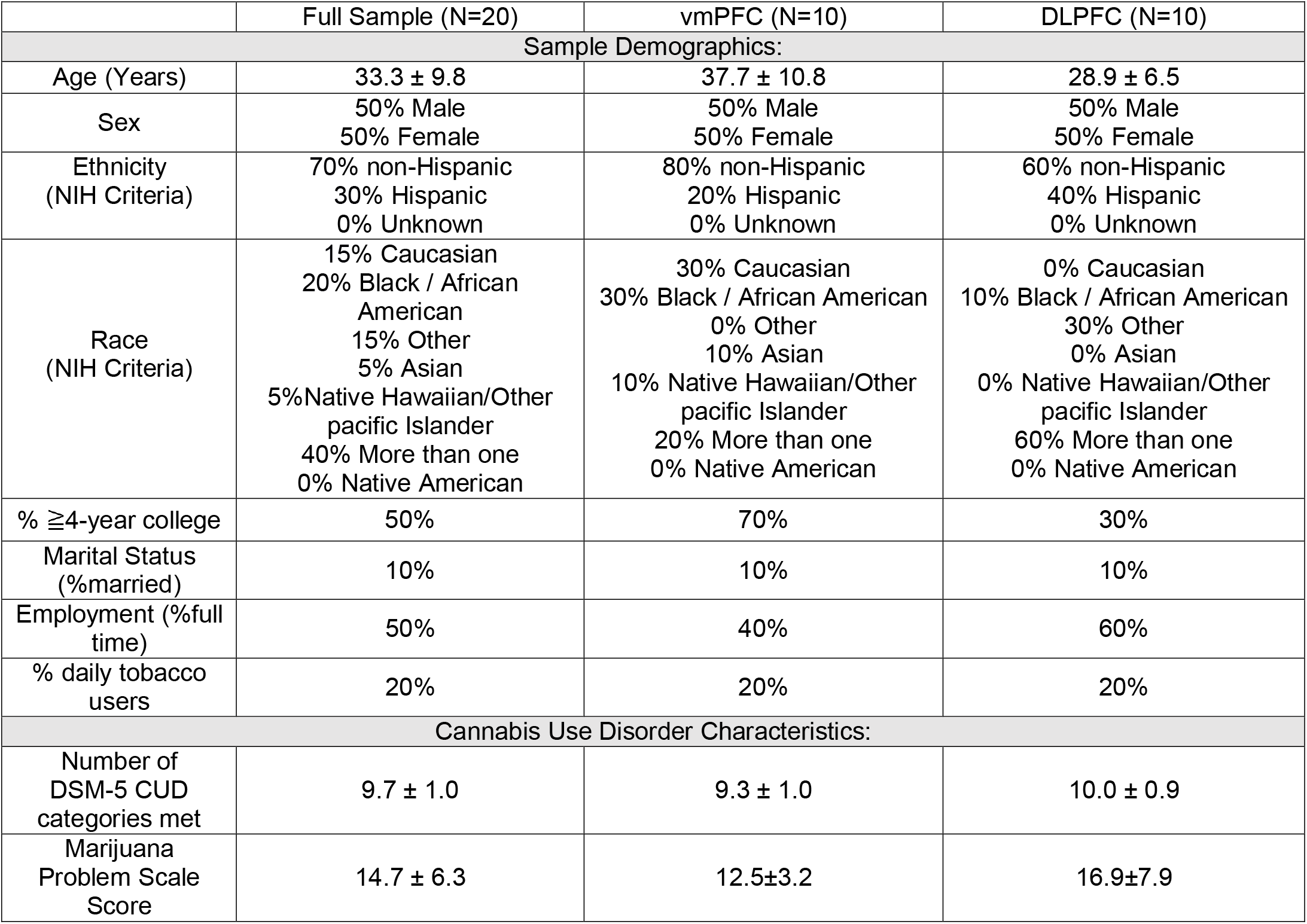
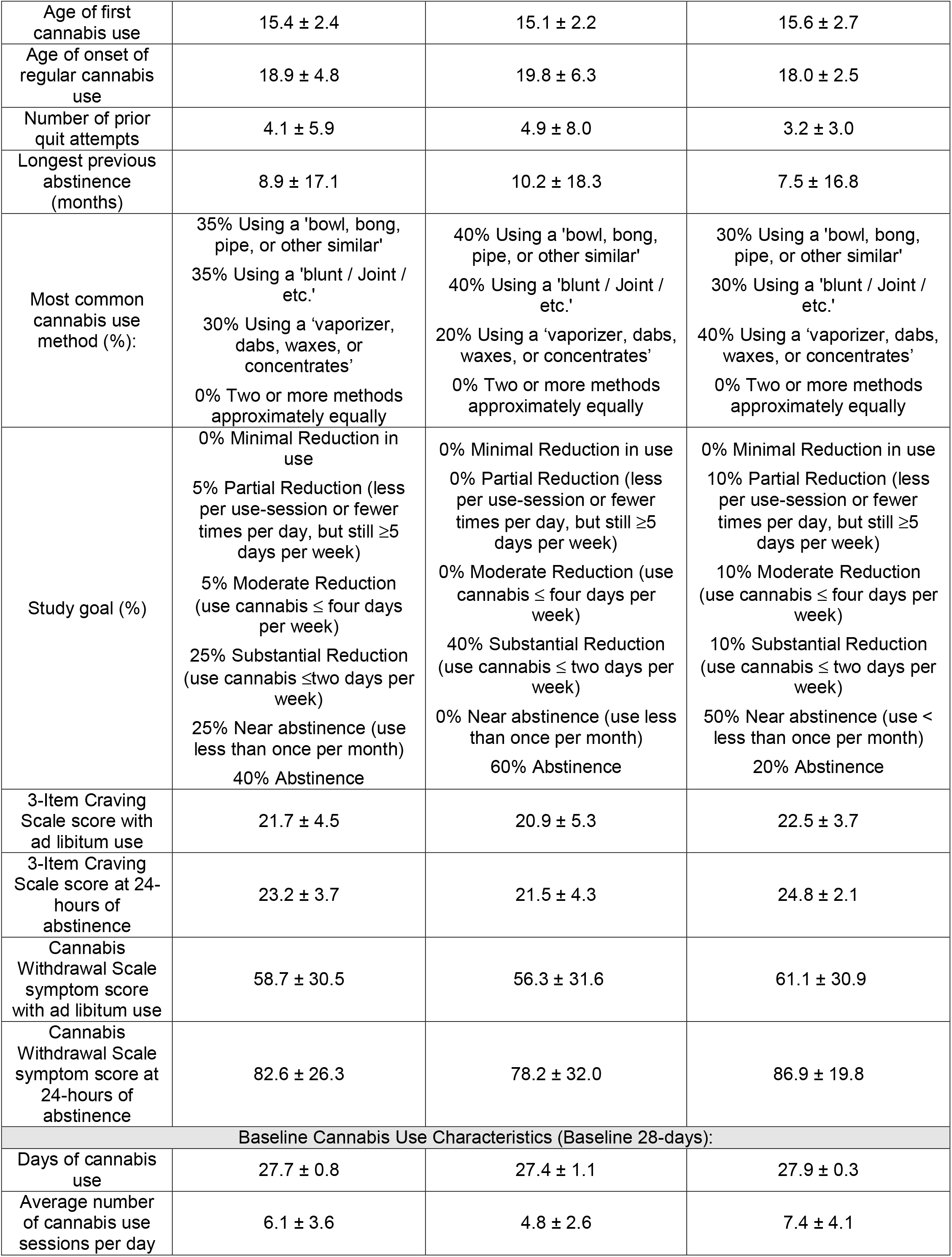
Baseline and demographic characteristics of the Intent to Treat (ITT) sample. All values are reported ± Standard Deviations. Cannabis use variables are reported for the 28-days prior to the screening and enrollment visit. DSM-5 CUD: Cannabis Use Disorder Criteria from the Diagnostic and Statistical Manual for Mental Disorders.

### Clinical Efficacy and adverse events (Figure 2, Supplemental table1)

A total of 43.1% of the weeks following the delivery of 16-sessions of vmPFC rTMS were self-reported as abstinent with 5 participants achieving 4 continuous weeks of abstinence in that time-period. There was no reported abstinence in the DLPFC group. Days-per-week and use-sessions-per-week both decreased from the baseline period to the follow-up period in both the vmPFC and DLPFC groups. There were significant decreases in days-per-week of cannabis use in both groups with large effect sizes (DLPFC: Cohens D = -3.1, *p*=0.01; vmPFC: Cohens D = -7.9, *p*<0.001). Similarly, both groups had a significant decrease in the number of use-sessions-per-week between baseline and follow-up periods (DLPFC: Cohens D = -0.88, *p*<0.001; vmPFC: Cohens D = -3.4, *p*<0.001). Quality of life also increased in both groups between the screening visit and 6-week follow-up (T(13)=-3.4, p=0.005, Cohens D=0.83). Adverse events were all mild and expected (Supplemental Table 1).

**Figure 1:**
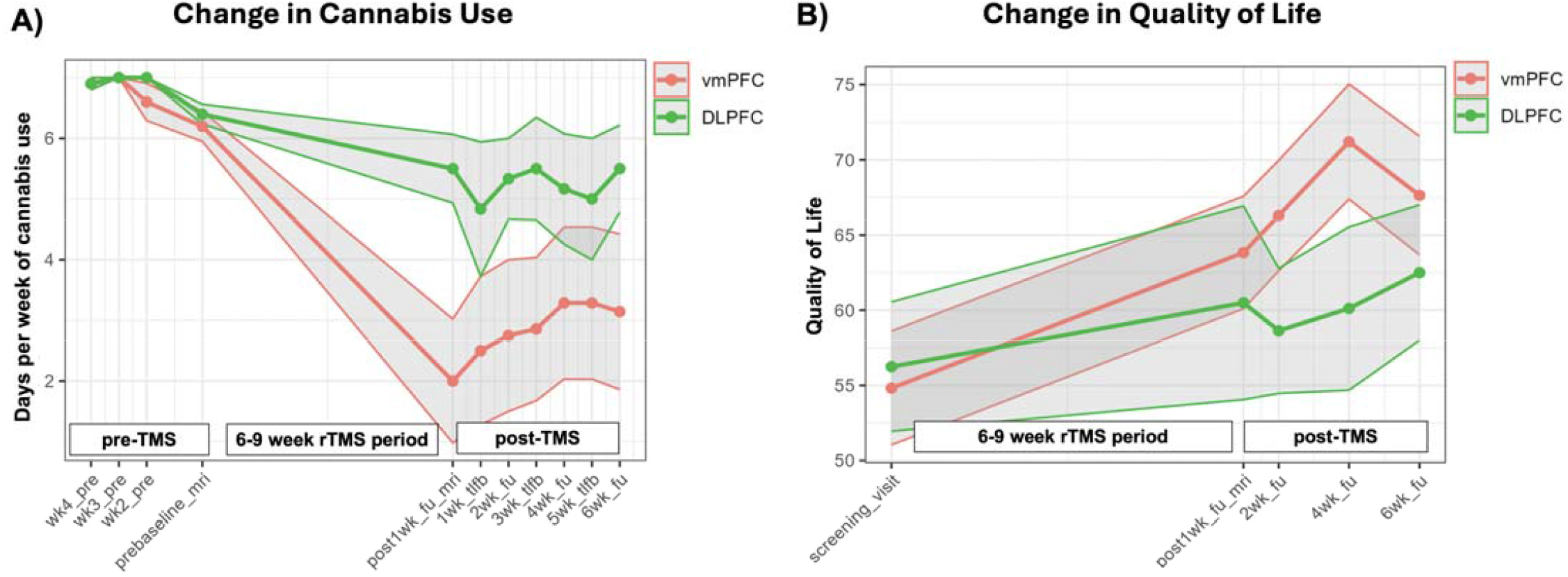
Change in days-per-week of cannabis use and quality of life following vmPFC and LDLPFC rTMS. Clinical outcomes were assessed via days-per-week of use (A) and quality of life (B), and are both presented longitudinally as average ± standard error of the mean (grey shaded area). Days-per-week of cannabis use was significantly reduced following both vmPFC (red) and DLPFC (green) rTMS (vmPFC: Cohens D = -7.9, *p*<0.001; DLPFC: Cohens D = -3.1, *p*<0.001;). Quality of life was significantly improved following both vmPFC and DLPFC rTMS (T(13)= 3.4, p=0.005, Cohens D= 0.83). wk=week; tlfb= Time-Line Follow-Back; fu=Follow up; vmPFC = ventromedial prefrontal cortex; DLPFC = dorsolateral prefrontal cortex.

**Figure 2:**
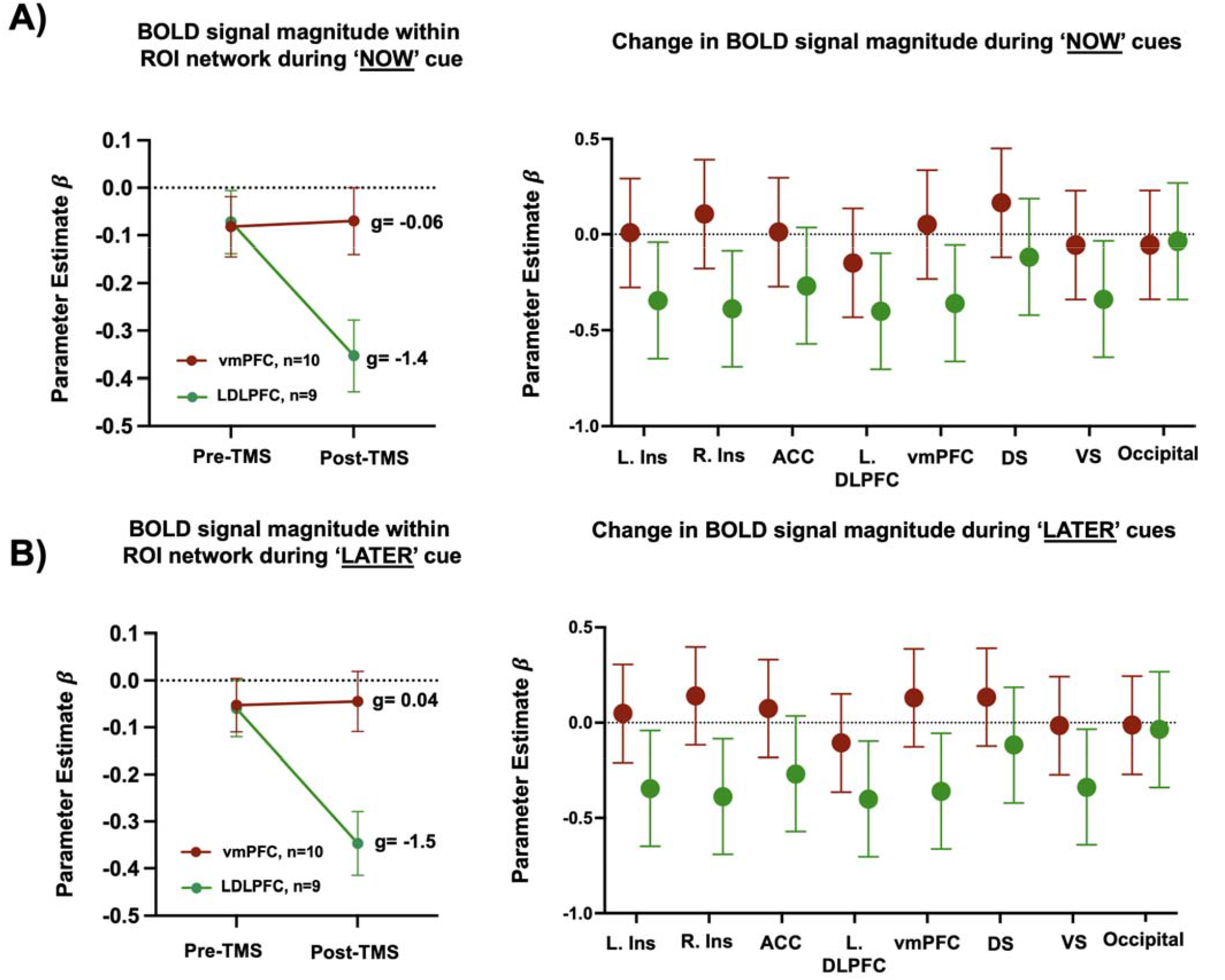
Change in fMRI BOLD signal magnitude in response to ‘Now’ and ‘Later’ cues following vmPFC and LDLPFC rTMS. A) Left: There was a large effect-size reduction (Hedge’s g) in BOLD signal magnitude in response to ‘Now’ cues following DLPFC rTMS (green) within the entire network of ROIs (left). Right: average change to BOLD signal magnitude within each ROI in the network. B) There was a large effect size reduction in response to ‘Later’ following DLPFC stimulation within the ROI network (left). Right: average change to BOLD signal magnitude within each ROI in the network. L=left, R=right, ACC=anterior cingulate cortex, DLPFC = dorsolateral prefrontal cortex, vmPFC = ventromedial prefrontal cortex, DS = dorsal striatum, VS = ventral striatum.

Craving scores decreased in both groups over time; however, the decrease did not significantly differ between the groups (p=0.21). The k-value of the $1000 delayed-discounting did not significantly decrease in either group, nor was the between group effects significantly different, though numerically there were between group differences in both assessments.

### Change in BOLD signal magnitude in response to ‘Now’ and ‘Later’ cues (Figure 2)

There was a significant main effect of treatment (F_1,271_=3.99, p=0.05), time (F_1,271_=3.86, p=0.05) and a time * treatment interaction (F_1,271_=4.61, p=0.03) when evaluating change in BOLD signal in response to ‘Now’ cues across the network of ROIs. Similarly, there was a significant main effect of treatment (F_1,271_=6.14, p=0.01), time (F_1,271_=4.95, p=0.03) and a time*treatment interaction (F_1,271_=5,55, p=0.02) when evaluating change in BOLD signal in response to ‘Later’ cues across the ROI network.

When ROI data were averaged across the entire network of regions, there was a greater reduction in BOLD signal magnitude in response to ‘Now’ and ‘Later’ cues following DLPFC rTMS (Now: vmPFC, Hedge’s g=-0.06; DLPFC, Hedge’s g=-1.4; Later: vmPFC: Hedge’s g=0.04; DLPFC, Hedge’s g=-1.53). DLPFC rTMS yielded medium to large effect-size reductions BOLD signal magnitude in response to ‘Now’ and ‘Later’ cues within all tested ROIs other than the occipital cortex, with the largest reductions occurring within the insula (Hedge’s g=-0.87), the striatum (Hedge’s g=-0.75) and the MPFC (Hedge’s g=-0.89). vmPFC rTMS produced a medium effect size increase in DLPFC reactivity to ‘Now’ and ‘Later’ cues (Hedge’s g=0.49 – 0.50) (Figure 2, Figure 4).

**Figure 3:**
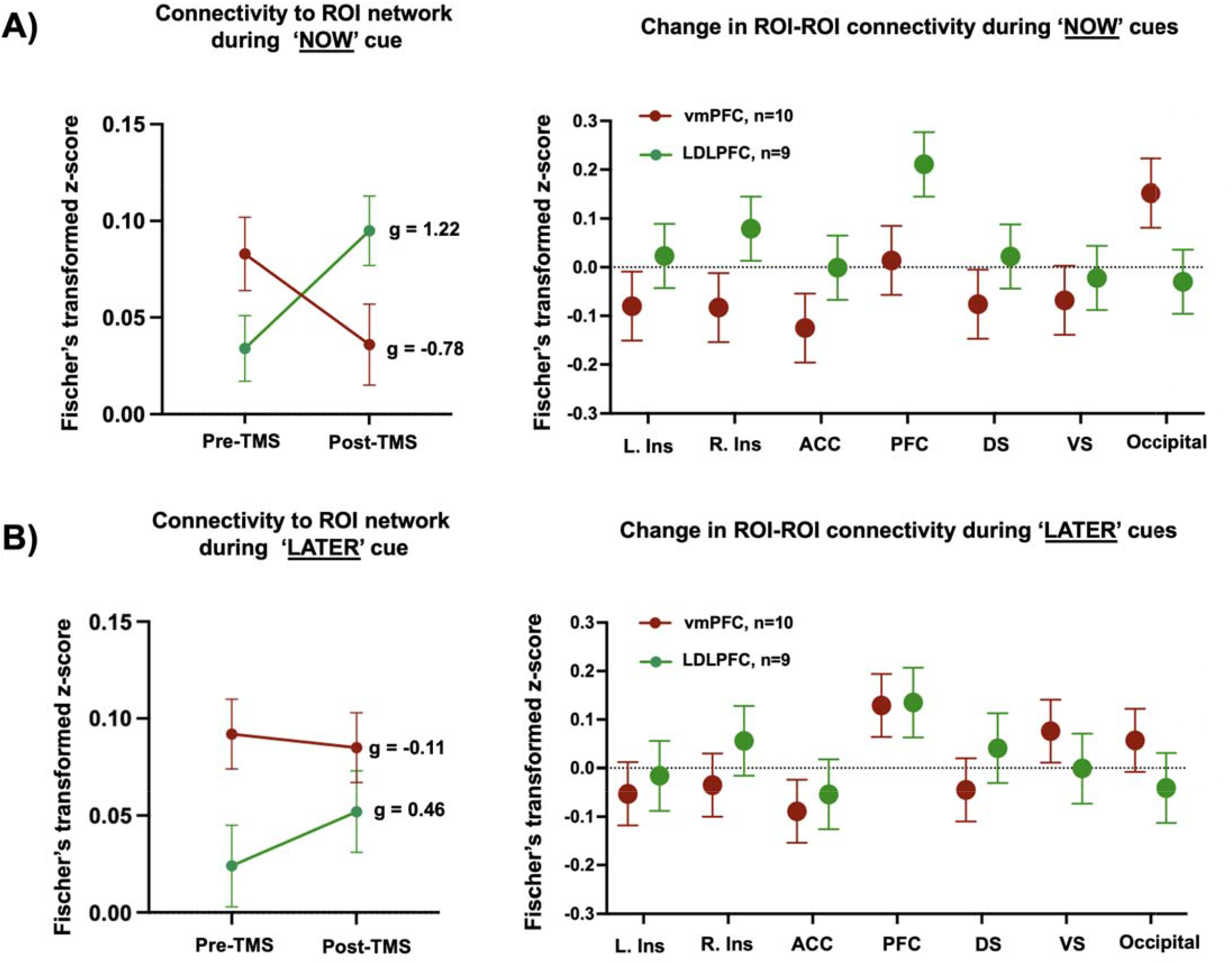
Change in ‘Now’ and ‘Later’ cue-induced functional connectivity following vmPFC and LDLPFC rTMS. A) There was a large effect-size increase (Hedge’s g) in functional connectivity in response to ‘Now’ cues following DLPFC rTMS (green) and a large effect-size reduction following vmPFC (red) stimulation (left). Right: average change to functional connectivity to each ROI in the network. ‘PFC’ represents connectivity to the cortical region that was not stimulated (e.g. vmPFC connectivity to the DLPFC in the vmPFC rTMS group; DLPFC connectivity to the vmPFC in the DLPFC connectivity group. B) There was a moderate effect size increase in functional connectivity in response to ‘Later’ cues following vmPFC stimulation (left). Right: average change to functional connectivity to each ROI in the network. L=left, R=right, ACC=anterior cingulate cortex, PFC = prefrontal cortex, DS = dorsal striatum, VS = ventral striatum.

**Figure 4:**
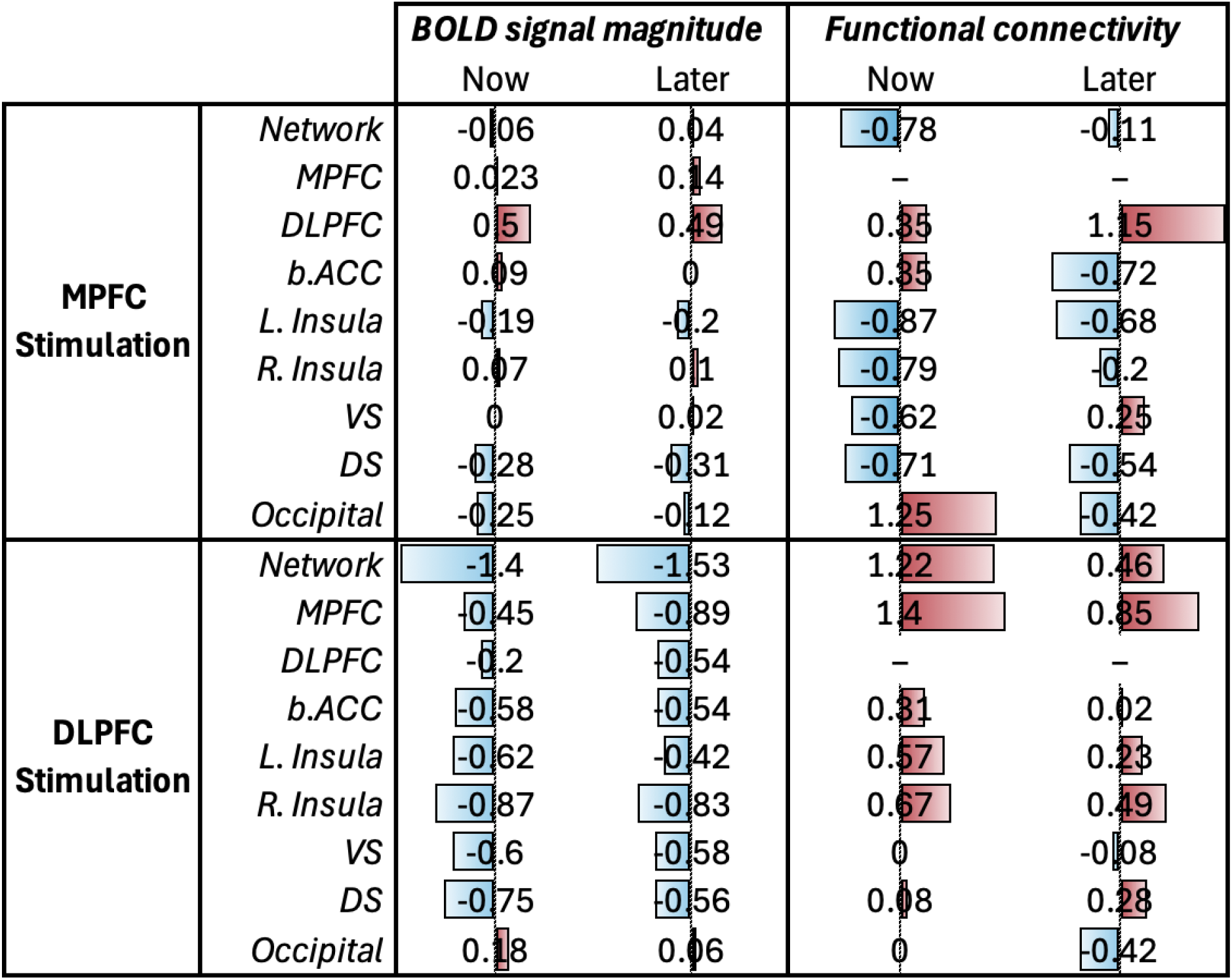
Summary of effect size changes in fMRI BOLD signal and functional connectivity following vmPFC and LDLPFC rTMS. Effect size change (Hedge’s g) on BOLD signal magnitude and functional connectivity following vmPFC and DLPFC stimulation. Relative increases and decreases in value are represented in red and blue, respectively. The length of each bar reflects the relative magnitude of the effect size.

### Change in functional connectivity during ‘Now’ and ‘Later’ cues (Figure 3)

There was a significant time*treatment interaction (F_1,244_=8.67, p=0.004) when evaluating change in functional connectivity during ‘Now’ cues and a main effect of treatment (F_1,244_=7.2, p=0.008) when change in ‘Later’ cues between the stimulation site and the network of ROIs.

When connectivity values were averaged across the network of regions, there was a large effect size decrease in connectivity following vmPFC rTMS (Hedge’s g=-0.78) and an increase in connectivity (Hedge’s g=1.22) following LDLPFC rTMS during ‘Now’ cues. There was a medium effect size increase in response to ‘Later’ cues following LDLPFC rTMS (LDLFPC: Hedge’s g= 0.46). vmPFC rTMS generally reduced functional connectivity during ‘Now’ and ‘Later’ cues, with the greatest reductions occurring between the vmPFC and the insula (Hedge’s g=-0.87), the striatum (Hedge’s g=-0.71), and the bilateral ACC (Hedge’s g=-0.72). There was a large effect size increase in functional connectivity between the vmPFC and LDLPFC following vmPFC rTMS (Hedge’s g=1.15) during ‘Later’ cues specifically. LDLPFC rTMS broadly increased functional connectivity between the LDLPFC and the insula (Hedge’s g=0.67) and the vmPFC (Hedge’s g=0.85) (Figures 3-4).

## Discussion

In this small proof of concept trial, we demonstrated the feasibility of this more sophisticated rTMS treatment paradigm for CUD as assessed by a strong retention rate (and low scores on the PeRBA), preliminary treatment efficacy for both treatment targets, and engagement of both behavioral and functional neuroimaging targets. In composite, these findings lay the foundation for further larger trials targeting the vmPFC and LDLFPC in the CUD population. We briefly discuss each of our findings in the following paragraphs as well as emphasize the various strengths and limitations of this completed study. Of important note, as will be emphasized again, though promising as these early results are, this small trial offers only hypothesis generating data and will need further expansion in a larger, blinded trial for confirmation.

### Feasibility

We generally found this more sophisticated treatment paradigm feasible to deliver. The retention rate of 80% was higher than the retention rate of our previous rTMS for CUD trial^9^, and our PeRBA scores were low and comparable to the other addictions-related trial that recorded them^35^. We previously reported the feasibility of generating functional rTMS targets using the ROC-task^20^ and found that we were able to generate robust targets for both the DLPFC and vmPFC in all participants who were able to complete MRI (19 out of 20 participants).

### Efficacy

In this open-label study we found significant decreases in days-per-week and use-sessions-per-week of cannabis between pre-treatment and post-treatment time-periods. We further observed a high rate of abstinence in the vmPFC group with five out of ten participants maintaining 4-weeks of continuous abstinence. We further found significant improvements in quality of life in both groups. Though there were numerically stronger reductions in cannabis use following vmPFC stimulation, any conclusions drawn from this observation are complicated by the relatively small sample size and the pre-existing differences in clinical presentation between groups. For example, the LDLPFC rTMS group entered the trial with more modest reduction goals and greater CUD severity scores, which may have limited treatment efficacy in this group. It is noteworthy that we replicated the finding of our previous trial in the important variable of decreasing days-per-week of cannabis use (with a larger effect size than our initial trial), an outcome that has consistently shown to result in improved quality of life and reduced cannabis related problems, anxiety, and depressive symptoms^36,37^. In a future larger and blinded trial, it will be possible to demonstrate definitively whether there is clinical efficacy at either or both treatment targets and whether there are behavioral or imaging findings that will help differentiate who will respond best to each treatment target.

### Changes to BOLD signal magnitude and functional connectivity

Brain reactivity to drug cues, as measured both by BOLD signal magnitude and functional connectivity, is a potentially valuable biomarker of relapse among individuals with substance use disorders^38^. We found distinct effects of vmPFC and LDLPFC rTMS in changing brain response to visual cannabis cues associated with the immediate, positive aspects of use (‘Now’) and the long-term, negative consequences of use (‘Later’). Primarily, we observed that LDLPFC rTMS broadly reduced BOLD signal magnitude in response to both cue types and increased functional connectivity in response to these cues across the network of ROIs. Similar reductions in BOLD signal magnitude following therapeutic courses of LDLPFC rTMS (e.g. greater than 10 sessions) have been observed among other populations with Alcohol Use Disorder^39,40^. While speculative in nature, it is possible the relative increase in functional connectivity values following LDLPFC stimulation may reflect an overall synchrony in network deactivation – that is, the stimulation site and downstream regions reduce their activity in unison. Interestingly, following vmPFC stimulation, we primarily observed reductions in functional connectivity, but not BOLD signal magnitude in response to cannabis cues. This finding has also been observed following rTMS for patients with Alcohol Use Disorder^33^. Notably, other published work has detected initial reductions in connectivity following vmPFC stimulation, which were subsequently followed by BOLD signal magnitude reductions months after treatment^33,41^. While again speculative in nature, this growing body of literature may support an overall desynchronization effect of vmPFC on brain reactivity to cues, specifically among regions involved in limbic and salience-based processes. Last, following both stimulation protocols, we observed a similar, large degree of increased connectivity between the vmPFC and LDLPFC. Several groups have documented that the degree of connectivity between these cortical regions is associated with reduced delayed discounting and greater self-control scores. Although underpowered to test whether this neural change mediated our clinical results, we highlight this as a worthwhile area of future exploration.

### A path forward – precision rTMS and individualization of cortical targets for rTMS in CUD and Substance Use Disorders

This study made two experimental advances that the rTMS field for addiction may consider incorporating moving forward to improve clinical efficacy: 1) an expanded number of rTMS sessions, consistent with field standards for rTMS in MDD and other disorders. For example, the rTMS for MDD literature consistently suggests that there is a dose-response effect for number of delivered treatments and remission rates^15,42–44^. A similar result is found in the single addiction trial that explores dose^35^. And, 2) individualization in rTMS targeting based on task-based fMRI signal – though scalp-based measurement targeting for rTMS is a valid method, and represents the clinical-backbone of targeting, an increasing sbody of literature suggests that anatomical and functional regions do not always overlap^45^, that treatment response in depression may be mediated by engaging sub-sections of the DLPFC connected to a deep brain structure (the sgACC)^13^, and more precise targeting of this deep brain structure using functional Magnetic Resonance Imaging (fMRI) may result in a larger antidepressant effect^14–16^. A recent study in addictions using task-based fMRI also demonstrated that rTMS delivered closer to an fMRI overlap location results in increased efficacy^17^. Taken together, the use of an fMRI guided rTMS targeting may be useful in determining which brain region to stimulate. The Regulation of Craving task, for example, consistently evokes activation in the vmPFC and DLPFC. Measuring the extent of brain reactivity within each target may be a useful way to select between these predominate cortical targets on an individual basis.

There were, of course, several limitations which included a relatively small sample size, the lack of a control group, and the completion of the trial at a single study-site. Due to these primary limitations, we were unable to draw extensive conclusions about which cortical target is more effective in treating CUD or perform any biotyping analyses. That said, we did generate a large set of effect sizes that may be used to power future comparative analyses between targets. We were also unable to find robust relationships between brain activity change and behavioral change within this small sample size.

In conclusion, in this preliminary trial, we found that 1) this experimental design was tolerable and feasible among this CUD population, 2) both rTMS strategies were preliminarily effective in reducing cannabis use days-per-week, and 3) rTMS delivered to the DLPFC and vmPFC had differential effects, with DLPFC rTMS reducing fMRI BOLD signal and increasing functional connectivity, and vmPFC rTMS primarily decreasing functional connectivity. These data may serve as a critical foundation to support future investigation into the continued development of rTMS for CUD.

## Data Availability

All data produced in the present study are available upon reasonable request to the authors

## Acknowledgements

The authors would like to thank the many contributors to this work including Lauren Crowe, Saron Hunegnaw, Noriah Johnson, Nour Dannawi, Ethan Makarewycz, Nolan Williams, and Leanne Williams.

## Supplementary Tables and Figures

**Supplemental Figure 1:**
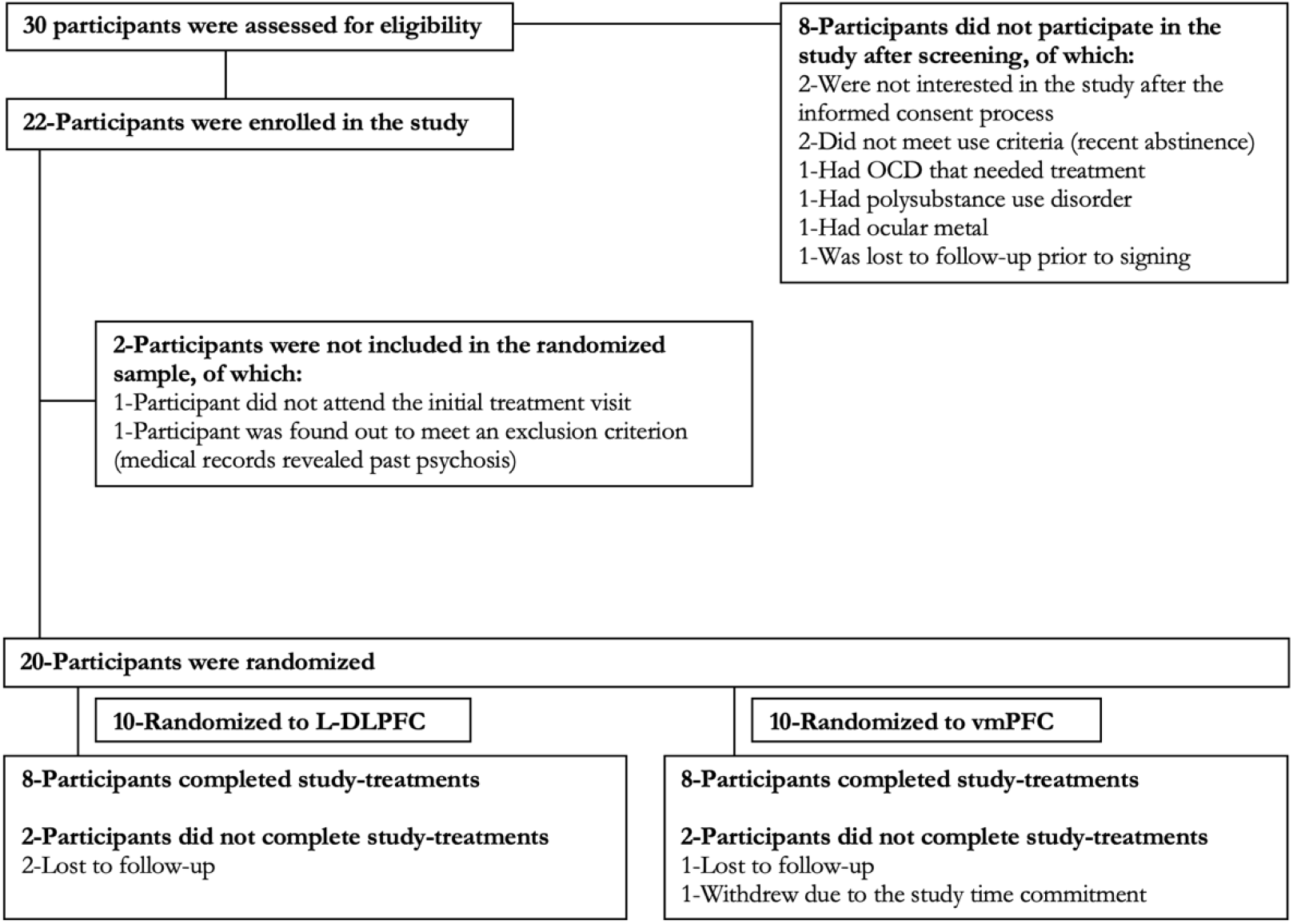
The figure below represents the CONSORT flow diagram for the study.

**Supplemental Figure 2:**
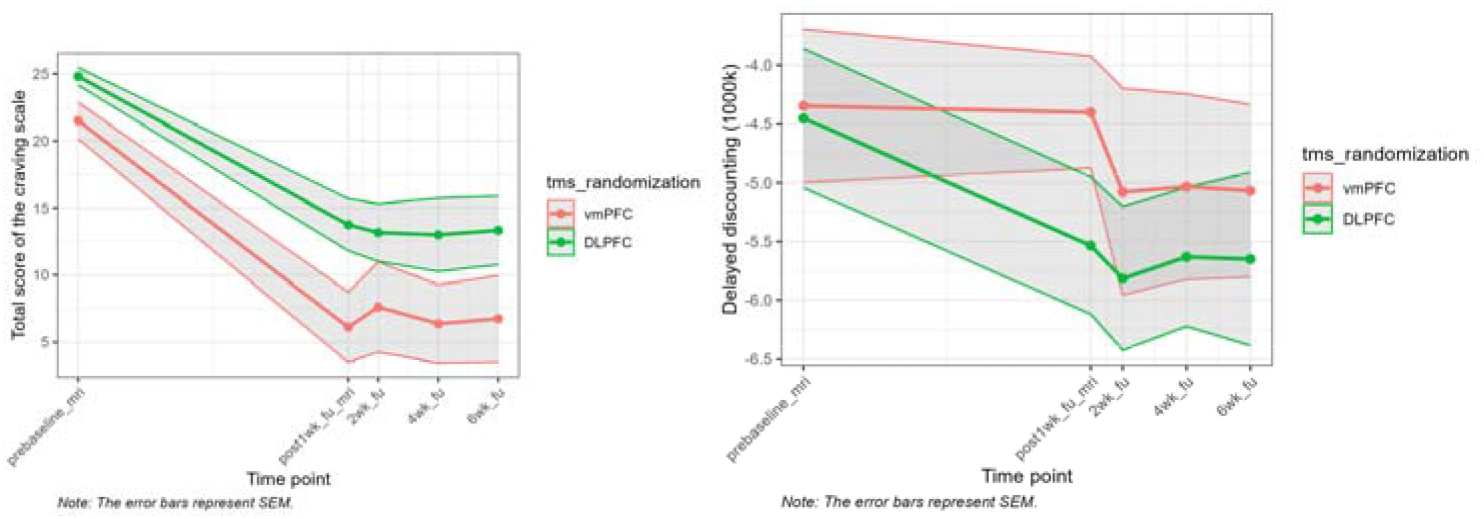
Craving, and Delayed Discounting.

**Supplemental Table 1:** Adverse Events (AE) between groups. The following AE table represents self-reported AE’s that were rated as probably not, possibly, probably, or definitely related to the study intervention. All AE’s were rated as mild in severity, and there were no serious AE’s.

|  | Total Sample, N, and<br>(% of ITT sample—<br>total / 20) | vmPFC condition, N,<br>and (% of ITT<br>sample—total / 10) | DLPFC condition, N,<br>and (% of ITT<br>sample—total / 10) |
| --- | --- | --- | --- |
| # of participants with Any<br>Adverse Events (%) | 6 (30%) | 3 (30%) | 3 (30%) |
| Total number of Adverse<br>events | 7 (35%) | 4 (40%) | 3 (30%) |
| Total number of serious AE's | 0 (0%) | 0 (0%) | 0 (0%) |
| Fatigue (%) | 3 (15%) | 1 (10%) | 2 (20%) |
| Headache (%) | 3 (15%) | 2 (20%) | 1 (10%) |
| Transient Vision Change (%) | 1 (5%) | 1 (10%) | 0 (0%) |

